# Epidemic Forecasting: Lessons Learned from the SARS-CoV-2 Pandemic to Balance Accuracy, Feasibility, and Impact

**DOI:** 10.1101/2025.11.03.25339385

**Authors:** Thomas Ferté, Vincent Thevenet, Xavier Hinaut, Pierrick Legrand, Dan Dutartre, Romain Griffier, Viannet Jouhet, Boris P. Hejblum, Rodolphe Thiébaut

## Abstract

The COVID-19 pandemic highlighted the importance of reliable, real-time hospital forecasting. At Bordeaux University Hospital, we developed models to predict SARS-CoV-2-related hospitalizations 14 days in advance using integrated data sources. We identified six key lessons to guide future epidemic response: (1) Multimodal data improves accuracy; (2) Simple baseline models are essential for benchmarking and building trust; (3) Model and metric choices must align with decision goals which often means prioritizing absolute over relative metrics and beginning with simple models; (4) Prediction intervals should be provided to communicate the uncertainty associated with forecasts; (5) Real-world constraints such as computational cost, maintainability, and required expertise should guide model selection; (6) Forecasts must be contextualized and communicated carefully to policymakers. We advocate for a systems-level forecasting approach that balances accuracy, feasibility, and impact.

## 1 Introduction

The COVID-19 pandemic, caused by SARS-CoV-2, emerged in late 2019 and quickly became a global health crisis (1, 2). Countries implemented enhanced surveillance, non-pharmaceutical interventions (e.g., lockdowns, curfews, school closures), and mass vaccination (3–6). Hospitals faced unprecedented pressure, prompting measures such as postponing elective procedures (7).

Anticipating epidemic trajectories was a major challenge, affecting timely and efficient care (8). Short-term forecasting models emerged but were limited by methodological heterogeneity, lack of standardization, and variable operational utility. Approaches ranged from simple extrapolation to advanced machine learning, often without clear guidance on comparative advantages (9, 10). At Bordeaux University Hospital (BUH), our multidisciplinary team developed a real-time forecasting model using penalized linear regression on hospital, public health, and weather data (11), later exploring machine learning and neural networks to improve predictions (12–14) . Beyond algorithmic performance, we faced practical challenges including forecast reliability, evaluation metrics, and communication with decision-makers.

This article provides an experience-based perspective on developing and deploying real-time epidemic forecasts, highlighting practical challenges, key decisions, and lessons for more actionable forecasting. Section 2 introduces the setting and models, followed by insights on data quality (Section 3), baseline models (Section 4), modeling approaches (Section 5), prediction uncertainty (Section 6), operational constraints (Section 7) and communication with policymakers (Section 8).

## 2 Methods

We illustrate our discussion using forecasting efforts during the COVID-19 epidemic at BUH, aiming to predict daily SARS-CoV-2 hospitalizations at a 14-day horizon. We chose a 14-day horizon to give the hospital enough time to respond while keeping forecasts practical (Section 4.2). Models were trained and evaluated every two days using integrated data from BUH’s Electronic Health Records, public COVID-19 statistics, weather, and vaccination coverage (15–17), spanning May 16, 2020, to January 17, 2022. The dataset included 409 predictors over 586 days. We compared three models:

- **Elastic net (ENet):** A simple, interpretable statistical model, easy to implement.
- **XGBoost:** A widely used tree-based machine learning algorithm for time series forecasting (9, 18).
- **Reservoir Computing (RC):** A neural network approach previously shown to perform best in our benchmarks (12) . RC uses a randomly initialized recurrent “reservoir” layer, training only the output layer to capture complex temporal dependencies efficiently (19–22). Hyperparameters were optimized using a Genetic Algorithm (12) . RC has been applied in diverse domains, including epidemic modeling, language processing, and financial forecasting (23–28).

Heavier models such as Transformers showed poor performance in this context (12) and were excluded. Additional methodological details, including preprocessing, model specifications, hyperparameter ranges, and feature selection, are provided in Supplementary Materials.

## 3 Key Point 1: Choosing the Right Data Sources

### 3.1 Multi-modal Data Integration

Comito et al. (9) reviewed SARS-CoV-2 forecasting features, including daily cases, deaths, recoveries, vaccination rates, and phone calls. They found no consensus on which features should be prioritized, likely due to local epidemic variability and prediction model differences. Similarly, Cramer et al. (29) observed no consistent benefit from using all available data sources in their benchmark. Nevertheless, most effective models integrate diverse data types.

**Weather.** Environmental factors like temperature and humidity influence viral spread (30, 31). Roumagnac et al. (31) proposed the IPTCC index, which scores transmission potential from 0 to 100 to capture this phenomenon. In our work, incorporating weather into 14-day forecasts improved accuracy: Median Absolute Error (MdAE) for Enet regression dropped from 17.21 to 11.63 (11), and RC Mean Absolute Error (MAE) decreased from 15.67 to 15.27 (12) .

**Electronic Health Records (EHRs).** Hospital data warehouses enable epidemic forecasting from real-time EHR streams, predicting admissions within hours or weekly visit volumes (11, 32, 33) . Aggregated EHR data in our models reduced 14-day forecast MdAE from 11.63 to 9.48 hospitalizations (11) .

**Mobility.** Mobility data from platforms such as Facebook, Google, and Baidu were used during the pandemic to capture population movement patterns. Several studies demonstrated their utility in improving forecast accuracy (34–36).

**Digital Surveillance.** Digital epidemiology tools, such as Google Trends and social media platforms, have been explored for epidemiological surveillance (37). However, findings in the context of COVID-19 have been mixed, due to the strong influence of media coverage and public announcements on search behavior, as well as variable lag times between symptom searches and case incidence (38, 39).

**Wastewater-Based Surveillance (WBS).** WBS provides a non-invasive method to monitor both symptomatic and asymptomatic SARS-CoV-2 infections by quantifying viral load in sewage. Unlike traditional testing, it captures a broader, more representative population signal. Studies have highlighted its early-warning potential, although timely sample processing is critical for realtime forecasting (40–43).

**Data Validity and Standardization Across Data Sources.** Beyond obtaining accurate data, a major challenge is the lack of standardized definitions across sources. For example, reported COVID-19 hospitalizations may vary widely depending on institutional criteria, particularly regarding when a patient is considered no longer infectious, underscoring the need for harmonized reporting to enhance forecasting accuracy.

### 3.2 Data Timeliness and Updating Frequency

Latency between events and database registration affects forecasting. Hospital data warehouses often provide faster updates than public sources. At BUH, clinical data were available within one day, compared with four-day delays in traditional epidemiologic surveillance. This faster access improved 14-day hospitalization forecast MdAE by 2.60 points (11) .

## 4 Key Point 2: Setting a Baseline Model

### 4.1 A Not-So-Trivial Baseline

A simple yet informative baseline model can be derived from available data. In our context, this baseline forecasts 14-day hospitalizations by assuming the value remains unchanged, essentially following the last observation carried forward (LOCF) method.

Eventhough outperforming such a naive baseline may seem trivial, empirical evidence suggests otherwise. Cramer et al. (29) evaluated 28 models predicting incident COVID-19 deaths. Only 18 out of 28 models achieved a Mean Absolute Error equal to or lower than this simple baseline. Similarly, Bracher et al. (44) found that 6 out of 8 models failed to outperform the baseline when forecasting two-week cumulative cases in Germany.

### 4.2 Why is this Baseline Model Useful?

**Detecting ineffective models.** Models that fail to outperform the baseline add no predictive value and are unlikely to support decision-making. In our setting, Transformer models exhibited this behavior (12) .

**Defining forecast horizons.** Neither XGBoost nor RC models outperformed the baseline for 21-day forecasts, suggesting that this horizon may be out of reach. Focus should instead be placed on shorter horizons (e.g., 7or 14-day) (12) . This aligns with prior findings that even 14-day forecasts are challenging, with typical delays of about six days between observed slope shifts and corresponding prediction changes (11) .

**Assessing data sufficiency.** Comparing model performance to this baseline over time provides key insights. Panel A of Figure 1 shows MAE typically around 20 hospitalizations, an arguably acceptable error margin. However, Panel B indicates that the Mean Absolute Error relative to the baseline (MAEB, i.e. the mean absolute difference between model error and baseline error) was often above zero before March 2021, meaning models frequently performed worse than the baseline during the early epidemic phase. This suggests that early in the pandemic, no model offered predictive value beyond the last observed data point.

**Figure 1:**
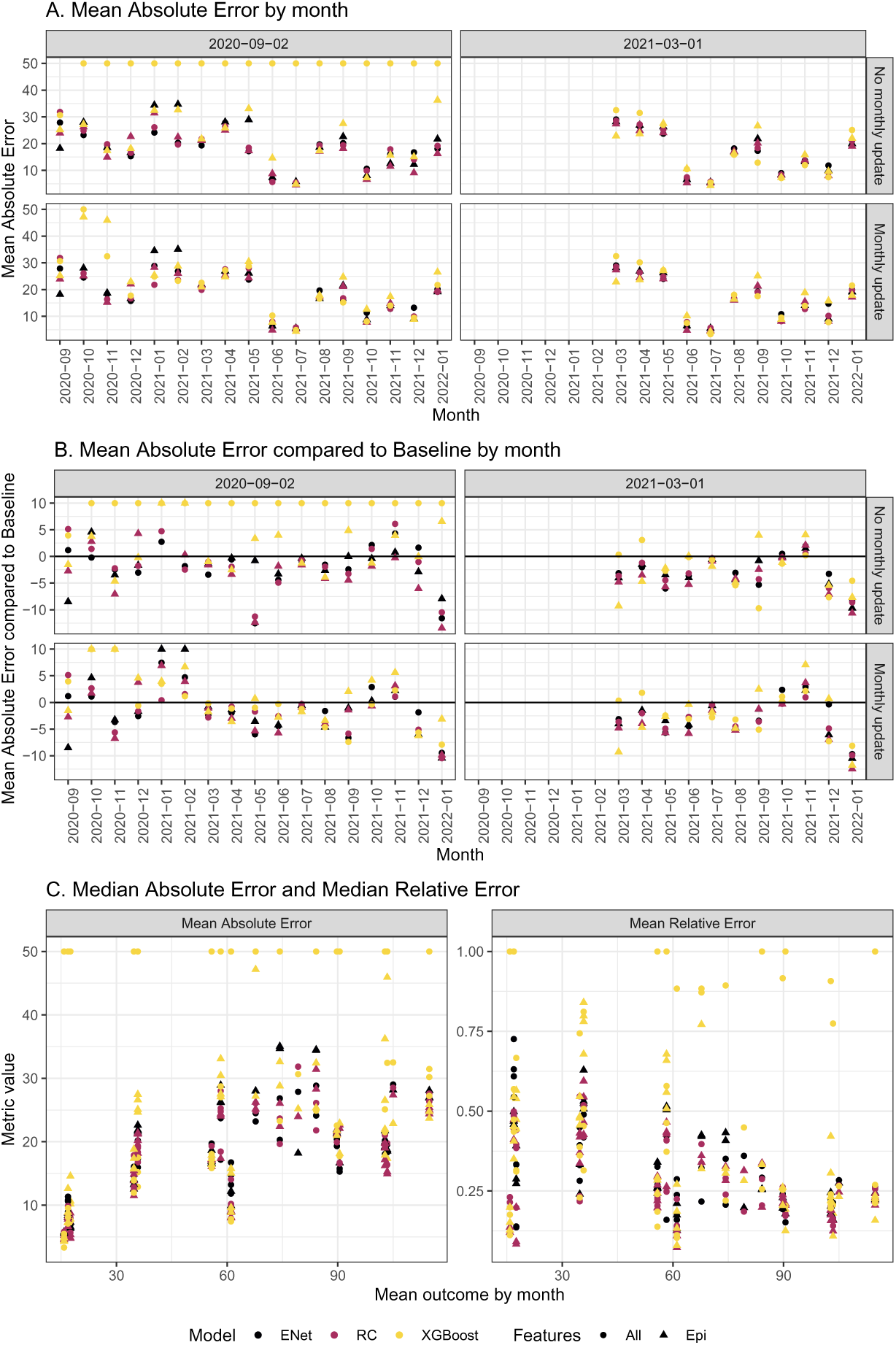
Panels A and B show MAE and MAEB by month. The first column shows models trained since 2020-09-02, and the second column shows models trained since 2021-03-01. The first row shows models without monthly updates, and the second row shows models with monthly updates. Panel C shows MAE and MAEB as a function of the mean outcome by month. MAE values above 50 were capped at 50, and MRE values above 1 were set to 1 for visualization purposes.

In summary, although straightforward to implement, this baseline model provides a key benchmark. It sets a minimum standard for forecasting performance that models must exceed, guiding model selection, informing forecast reliability assessments, and setting realistic expectations for predictive accuracy.

## 5 Key Point 3: Finding the best model

### 5.1 The right metric for the right task

Comito et al. (9) found 14 different metrics used across forecasting studies, with most reporting multiple metrics and few designating a primary one. Exceptions include Zheng et al. (45) and Gerlee et al. (46), who emphasized Mean Absolute Percentage Error (MAPE), though the rationale for prioritizing it over absolute metrics like MAE is unclear.

Choice of metric matters because different metrics emphasize different performance aspects and affect model selection. Absolute metrics (e.g., MAE, MSE) measure deviations in raw hospitalization counts, while relative metrics (e.g., MAPE, Laplacian Error) scale errors by observed values, effectively downweighting high-incidence deviations.

Our goal was to forecast hospital occupancy during epidemic peaks, when underestimating hospitalizations risks capacity strain and delayed care. Metrics sensitive to peak errors are therefore preferable. Figure 1 shows that MAE rises with hospitalization counts, capturing large deviations during peaks, whereas

Median Relative Error (MRE) down-weights them. For anticipating hospital surges, MAE is more appropriate.

In summary, aligning metrics with forecasting objectives ensures model evaluation supports real-world decision-making.

### 5.2 Data preparation

Preprocessing is often beneficial before feeding data into a model. In the context of COVID-19 forecasting, three key aspects are smoothing, computing derivatives, and transforming outcomes.

**Smoothing.** Because COVID-19 data are highly variable over time, smoothing can facilitate model learning. In Ferté et al. (11), local polynomial smoothing reduced the MdAE by 2.47 hospitalizations compared to no smoothing. Paireau et al. (34) also showed that it outperformed a simple 7-day moving average.

**Derivatives.** Computing derivatives can also improve performance. In linear regression, derivatives reduced MdAE by 3.24 hospitalizations (11) . Even with more complex models such as RC, explicitly including derivatives reduced MAE by 3.47 hospitalizations (12), suggesting that even complex models do not always extract this information spontaneously.

**Outcome transformations.** Learning directly from raw outcomes is not always optimal. Alternatives include learning growth rates (34) or applying log-transformations (47), both based on the assumption of local exponential growth. In our work, we proposed learning the residuals of the LOCF baseline relative to hospitalizations rather than raw hospitalization counts, thereby simplifying the task (14) . In other words, we modeled the change in hospitalizations rather than their absolute values.

### 5.3 Statistical learning vs Machine Learning

Several articles have explored the distinctions and overlap between statistical learning and machine learning (48–50), including in the context of biopreparedness (51). Statistical learning is typically valued for its interpretability, simplicity, and speed, though it often relies on linear assumptions and struggles with high-dimensional data. In contrast, machine learning handles complex, non-linear patterns and high-dimensional inputs better, but is generally less interpretable and more computationally intensive.

This contrast is not absolute. Feature engineering can address some limitations of statistical models, whose interpretability also declines with more and correlated features. Meanwhile, machine learning has become more accessible thanks to modern software, and its computational demands are increasingly manageable.

The two paradigms also differ in approach: epidemiology-driven modeling emphasizes expert feature selection, while machine learning favors a more datadriven, holistic strategy. As Beam et al. (50) suggests, these approaches form a continuum, where more data often calls for more automated methods.

To examine these differences, we selected three representative methods: Enet for statistical modeling, XGBoost for tree-based machine learning, and RC for neural network-based learning. We then explore the following hypotheses: (1) machine learning requires more data than statistical models; (2) machine learning can outperform statistical models due to fewer assumptions and (3) expert-driven feature selection outperforms purely data-driven selection.

#### 5.3.1 Quantity of data needed

We first examined a setting with limited information (2020-09-02 to 2021-03-15, Table 1). We hypothesized that simpler models would outperform complex ones, which risk overfitting. Indeed, linear regression with expert-selected features achieved the lowest MAE. However, the MAEB metric showed only a marginal improvement over the LOCF baseline (0.56 hospitalizations), a clinically negligible difference (Figure 2). Overall, when data are limited, neither statistical nor machine learning models provide accurate forecasts.

**Figure 2:**
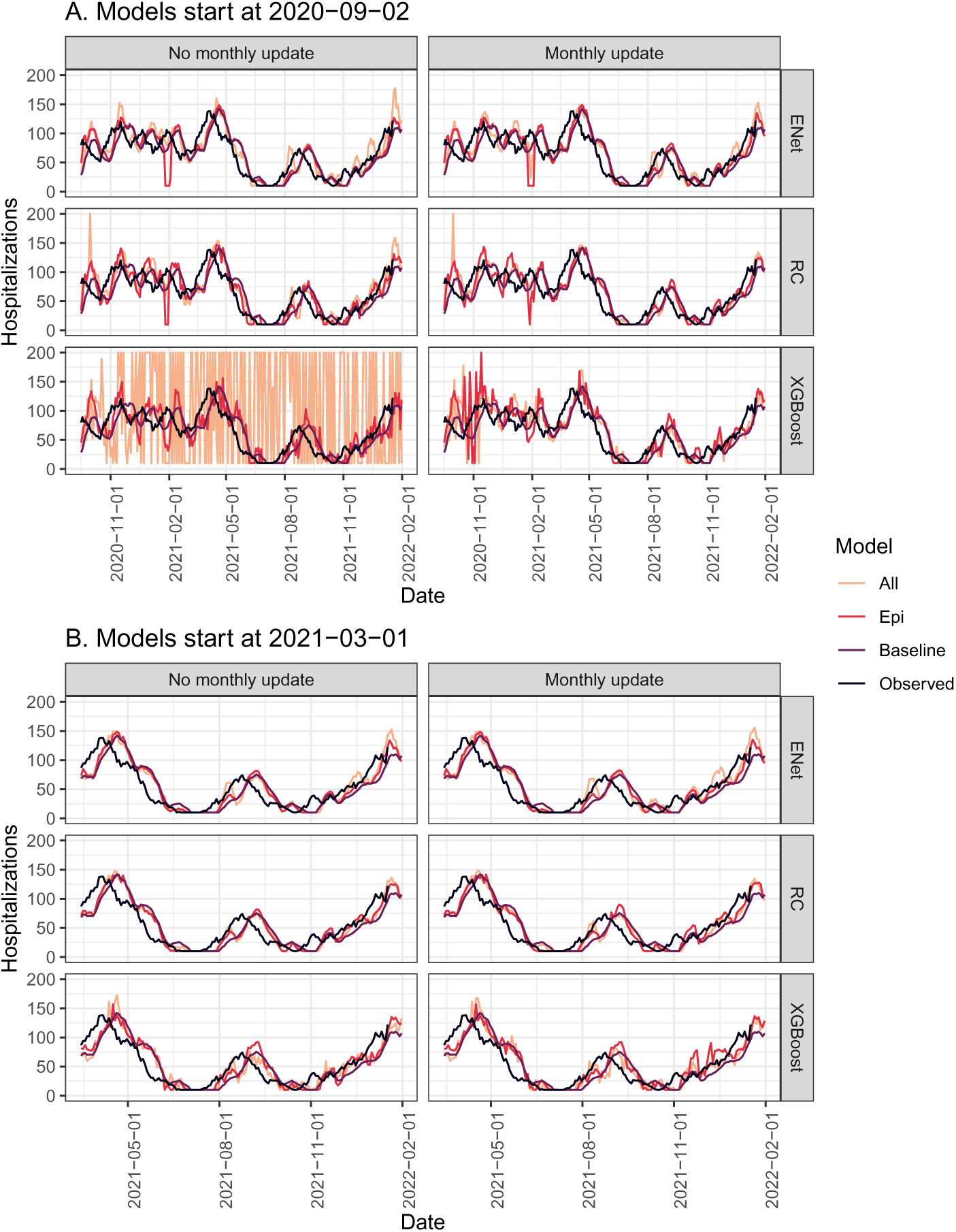
Forecasts generated by different models initialized at various start dates, with or without monthly hyperparameter updates, using either all available features (All) or a feature set selected based on a priori knowledge (Epi).

**Table 1:** Performance by algorithm for models initiated on 2020-09-02 and evaluated from 2020-09-02 to 2021-03-15.

| Model | Features | Update | MAE (± SD) | MAEB (± SD) |
| --- | --- | --- | --- | --- |
| Baseline | - | - | 21.03(±12.45) | 0.00(±0.00) |
| ENet | all | No | 20.47(±13.86) | -0.56(±14.04) |
| | all | Yes | 22.12(±15.73) | 1.09(±14.23) |
| | epi | No | 24.36(±20.26) | 3.33(±17.76) |
| | epi | Yes | 24.65(±20.54) | 3.61(±18.33) |
| RC | all | No | 21.54(±18.39) | 0.51(±19.52) |
| | all | Yes | 20.82(±18.42) | -0.21(±17.7) |
| | epi | No | 22.47(±18.41) | 1.44(±18.5) |
| | epi | Yes | 22.69(±16.69) | 1.66(±17.0) |
| XGBoost | all | No | 161.48(±253.34) | 140.44(±255.67) |
| | all | Yes | 28.72(±24.91) | 7.68(±23.19) |
| | epi | No | 24.95(±19.65) | 3.91(±19.25) |
| | epi | Yes | 31.43(±37.04) | 10.39(±36.03) |

#### 5.3.2 Models comparison

We then assessed whether machine learning outperforms statistical learning with more data (Table 2). All models exceeded the baseline by about 2.5 hospitalizations. XGBoost performed worse than both RC and ENet, while RC slightly outperformed ENet by less than one hospitalization.

**Table 2:** Performance by algorithm for models initiated on 2021-03-01 and evaluated from 2021-03-15 to 2022-01-17. Each algorithm was repeated three times; the table shows the median MAE and MAEB along with their minimum and maximum values.

| Model | Features | Update | MAE (min ; max) | MAEB (min ; max) |
| --- | --- | --- | --- | --- |
| Baseline | - | - | 18.59 (18.59 ; 18.59) | 0.00 (0.00 ; 0.00) |
| ENet | all | No | 15.83 (15.82 ; 15.87) | -2.76 (-2.77 ; -2.72) |
|  | all | Yes | 15.98 (15.9 ; 16.29) | -2.6 (-2.68 ; -2.29) |
|  | epi | No | 15.93 (15.86 ; 16) | -2.65 (-2.73 ; -2.58) |
|  | epi | Yes | 15.88 (15.87 ; 15.89) | -2.7 (-2.72 ; -2.7) |
| RC | all | No | 15.4 (15.24 ; 15.6) | -3.19 (-3.35 ; -2.99) |
|  | all | Yes | 15.63 (15.59 ; 15.65) | -2.96 (-3 ; -2.93) |
|  | epi | No | 15.16 (15.05 ; 17.45) | -3.43 (-3.53 ; -1.14) |
|  | epi | Yes | 15.1 (14.96 ; 15.46) | -3.48 (-3.62 ; -3.12) |
| XGBoost | all | No | 16.08 (15.98 ; 19.95) | -2.51 (-2.61 ; 1.36) |
|  | all | Yes | 16.29 (16.26 ; 16.78) | -2.29 (-2.33 ; -1.8) |
|  | epi | No | 16.44 (16.42 ; 16.97) | -2.15 (-2.17 ; -1.62) |
|  | epi | Yes | 16.99 (16.61 ; 17.32) | -1.59 (-1.97 ; -1.27) |

These results highlight that statistical learning remains competitive with machine learning approaches. Also, as expected, all machine learning approaches are not behaving the same and careful choice should be done among these approaches. In our setting, the last neural network layer of RC is a linear regression, making this method conceptually closer to Enet than to XGBoost. In other words, the question is not to choose between statistical and machine learning but the best method according to the context.

#### 5.3.3 Feature selection

We also evaluated how expert-driven feature selection compares to model-based selection using all available features. The expert-selected set included hospitalizations, positive RT-PCR cases, the proportion of positive RT-PCRs among individuals over 60, IPTCC, the cumulative number of first-dose vaccinations, and emergency department visits with “COVID-19” mentioned in the EHR. These variables were chosen by an epidemiologist based on prior knowledge before fitting the models.

As Table 2 shows, differences in MAE were under 0.5 hospitalizations across all algorithms. Enet was unaffected. RC showed a slight improvement with the expert-selected features, possibly due to the advantage of starting with a smaller, more focused input set before reservoir-based dimensional expansion. XGBoost performed slightly worse with the expert-selected features.

While the performance differences are modest, expert-driven feature selection may offer benefits from an implementation science perspective, such as improved interpretability and ease of adoption, as discussed in Section 7.

## 6 Key Point 4: Uncertainty quantification

Uncertainty quantification is essential for informed decision-making, especially in COVID-19 forecasting where epidemiological dynamics change rapidly. Estimating prediction intervals is challenging due to temporal correlations.

Initially, we applied a simple *±*40% range around predictions, which yielded only 51% empirical coverage—insufficient for epidemic forecasting (11) . We refined this approach, denoted as Ad hoc in Figure 3, by expanding intervals by *±x*%, calibrated to achieve 95% coverage using data from 2020-09-02 to 2021-03-01 as a calibration set.

**Figure 3:**
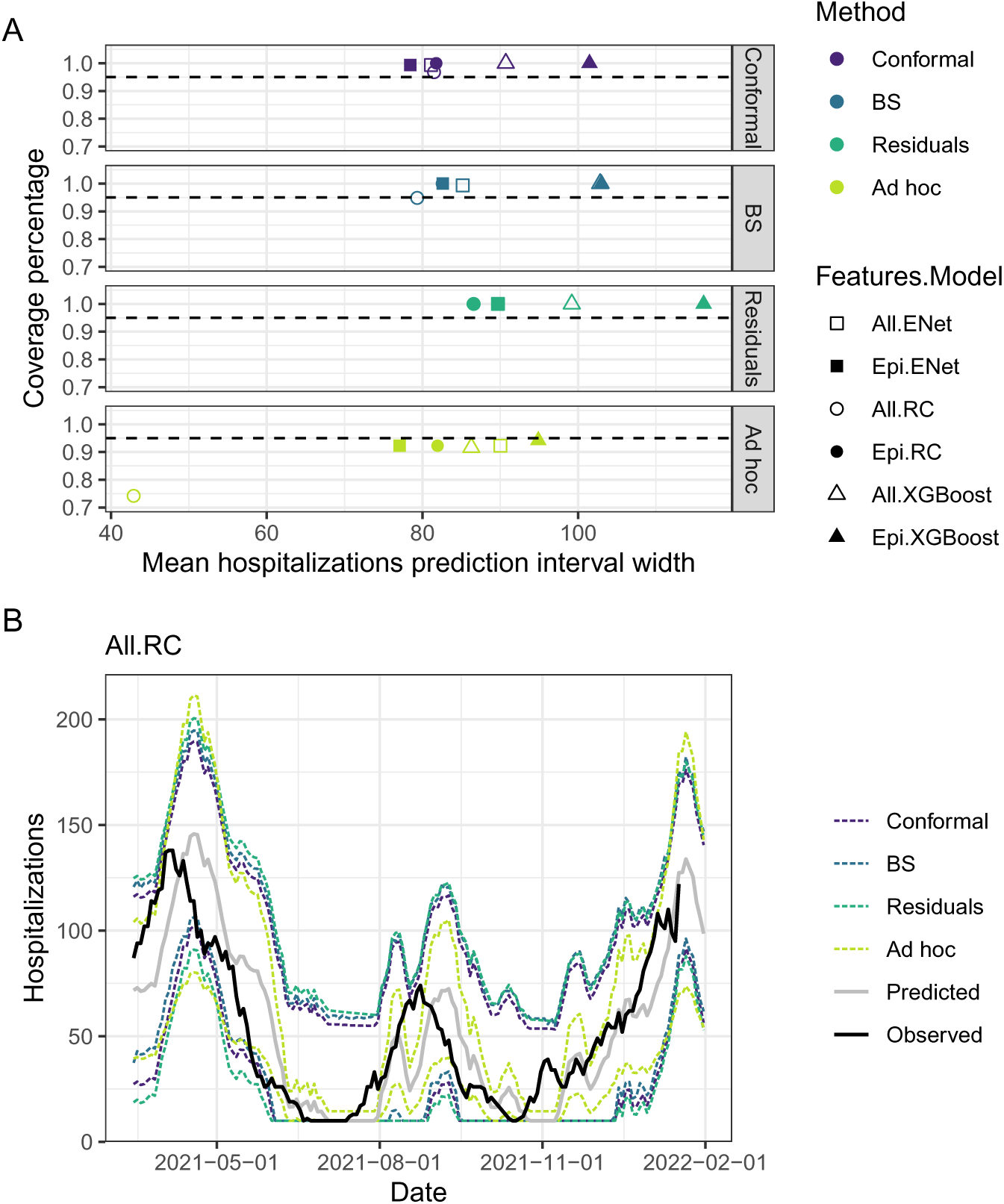
Estimating model uncertainty. A) Coverage percentage and width of 95% prediction intervals (PI); the dotted line marks the target coverage. The best method achieves coverage of 95% with the narrowest interval width. B) Prediction intervals for RC predictions using all features and different estimation methods. *Residuals* and *BS* estimate PIs from the distribution of past residuals (52). *Conformal* applies an adaptive method to iteratively widen or tighten the PIs (53). The *Ad hoc* method adds *±x*% to predictions based on residuals observed before 2021-03-01 (11) .

To further improve uncertainty estimation, we considered model-agnostic post hoc methods that leverage past residuals. In our setup, residuals from 2020-09-02 to 2021-03-01 form an initial calibration set, updated daily as new residuals become available, and are used to evaluate three additional approaches.

Two baseline methods are shown in Figure 3: *Residuals*, which assumes normality to compute 95% intervals, and *Bootstrap (BS)*, which samples 1,000 past residuals to construct empirical 95% quantiles (52). Both methods adapt slowly, as older residuals are weighted equally with more recent ones.

Conformal prediction (CP) addresses this limitation. Adaptive Conformal Inference (ACI) adjusts interval widths daily based on prior coverage, controlled by a hyperparameter *γ* (54). AgACI extends this approach by automatically tuning *γ* via online expert aggregation (53), implemented in the AdaptiveConformal R package (55).

Figure 3 summarizes the performance of these methods; additional details are in Supplementary 10.2. All approaches except the Ad hoc method reached the nominal 95% coverage, though some slightly over-covered. AgACI achieved the narrowest intervals, followed closely by BS, while Residuals underperformed, likely due to the normality assumption. The Ad hoc strategy failed to reach 95% coverage.

Looking ahead, BS and CP methods appear to be the most promising for future epidemic forecasting. CP is theoretically well-suited to handle abrupt distributional shifts, due to its online adaptive nature. This is a particularly valuable property in the context of rapidly evolving epidemics. However, in practice, our empirical results showed little difference in performance between CP and BS methods.

While RC achieved the highest forecasting performance, its advantage over ENet was modest, highlighting the need to consider criteria beyond predictive accuracy. Using the Consolidated Framework for Implementation Research (CFIR) (56), we assess model deployment across five implementation domains, emphasizing the Innovation domain.

### 6.1 Innovation

The Innovation domain in CFIR focuses on characteristics of the intervention, including Relative Advantage, Complexity, Cost, Design Quality and Packaging, and Trialability. Table 3 summarizes the three forecasting models on these dimensions.

**Table 3:** Comparison of ENet, XGBoost, and RC for the Innovation domain of the CFIR framework.

| Innovation | ENet | XGBoost | RC |
| --- | --- | --- | --- |
| Relative Advantage | ⚖️ Slightly lower performance | ✗ Lower performance | ✓ Best forecasting accuracy |
| Complexity | ✓ Simple and familiar | ⚖️ Standard libraries available | ✗ Less common, more complex due to genetic tuning |
| Cost | ✓ Low computational and human resource cost | ⚖️ Computational cost depends on feature set | ✗ Moderate computational cost, higher human effort |
| Design Quality & Packaging | ✓ Well-supported and easy to integrate into standard pipelines |  | ✗ Requires custom integration and maintenance for genetic optimization |
| Trialability | ✓ All models can be readily tested retrospectively with standard cross-validation frameworks |  |  |

**Table 4:** Hyperparameter ranges

| Model<sup>1</sup> | Hyperparameter | Range | Log<sup>2</sup> |
| --- | --- | --- | --- |
| XGBoost | n_estimators | 3;300 | False |
|  | max_depth | 5;100 | False |
|  | learning_rate | 1e-5;1 | True |
|  | subsample | 0;1 | False |
|  | colsample_bytree | 0;1 | False |
| ENet | ridge | 1e-10;1e5 | True |
|  | l1_ratio | 0;1 | False |
| RC | ridge | 1e-10;1e5 | True |
|  | spectral_radius | 1e-5;1e5 | True |
|  | leaking_rate | 1e-5;1 | True |
|  | input_scaling | 1e-5;1e5 | False |
|  | feature_selection<sup>3</sup> | y, n |  |

<sup>1</sup> ENet : Penalized linear regression, RC : Reservoir computing
<sup>2</sup> For features with *Log = True*, features are log-transformed before being processed by GA or RS
<sup>3</sup> One hyperparameter per feature

**Relative Advantage.** RC achieved the lowest mean absolute error, but its advantage over ENet was modest, raising the question of whether the performance gain justifies the added complexity.

**Complexity.** ENet is the simplest and widely familiar. XGBoost is also wellknown with robust libraries. RC, particularly with genetic algorithm tuning, is technically complex and less familiar to practitioners.

**Cost.** ENet has a clear advantage in terms of both computational and human resource costs, which remain low even at scale. XGBoost’s computational cost varies depending on the number of features and parameter settings, but its practical cost remains moderate. In contrast, RC with genetic optimization introduces moderate computational demands and requires additional human expertise for setup and maintenance.

**Design Quality and Packaging.** ENet and XGBoost are well-supported within existing data science ecosystems, offering straightforward implementation and mature libraries in both Python and R. RC, particularly when combined with genetic algorithms, is less seamlessly integrated. However, the ReservoirPy module provides a flexible Python implementation, and we recently developed the reservoirnet package to extend this accessibility to R users (14, 57) .

**Trialability.** All three models have high trialability, as they can be rigorously tested on retrospective data before daily deployment. We implemented an accumulative validation framework (58), where models were retrained daily on all available data to mimic real-time conditions, with hyperparameters re-optimized monthly to reflect realistic resource constraints. Full details are provided in Section 10.1.2 and Ferté et al. (12) .

Overall, this comparison highlights a trade-off between accuracy and ease of implementation. RC delivers highest performance, while ENet’s simplicity, low cost, and familiarity make it a strong alternative. In our setting, XGBoost offered no clear performance advantage to justify its added complexity.

### 6.2 Inner Setting

Implementation of forecasting models was enabled by a well-established hospital data warehouse and an existing daily dashboard reporting key indicators such as hospitalizations, confirmed cases, and ICU admissions, providing a strong foundation for integration (11) .

Computing capacity posed little barrier for lightweight models like ENet, though more resource-intensive methods such as RC with genetic algorithms could have been constrained.

Strong collaboration between the epidemiology–biostatistics and medical informatics teams, built through prior joint projects, fostered trust and a shared problem-solving culture, which supported timely implementation (59) .

### 6.3 Outer Setting

The SARS-CoV-2 pandemic was the main external driver, creating urgent demand for timely data to guide crisis response and mobilizing healthcare professionals.

While no direct environmental constraints affected our project, awareness is growing around the carbon footprint of data-intensive methods. As shown in Figure 4, ENet had the lowest emissions, XGBoost’s footprint increased with feature set size, and RC with genetic algorithms was most energy-intensive. The burden also depends on local energy mixes: France’s electricity (51.28 gCO_2_/kWh) is far less carbon-intensive than Germany’s (338.66) or the U.S.’s (423.94) (60).

**Figure 4:**
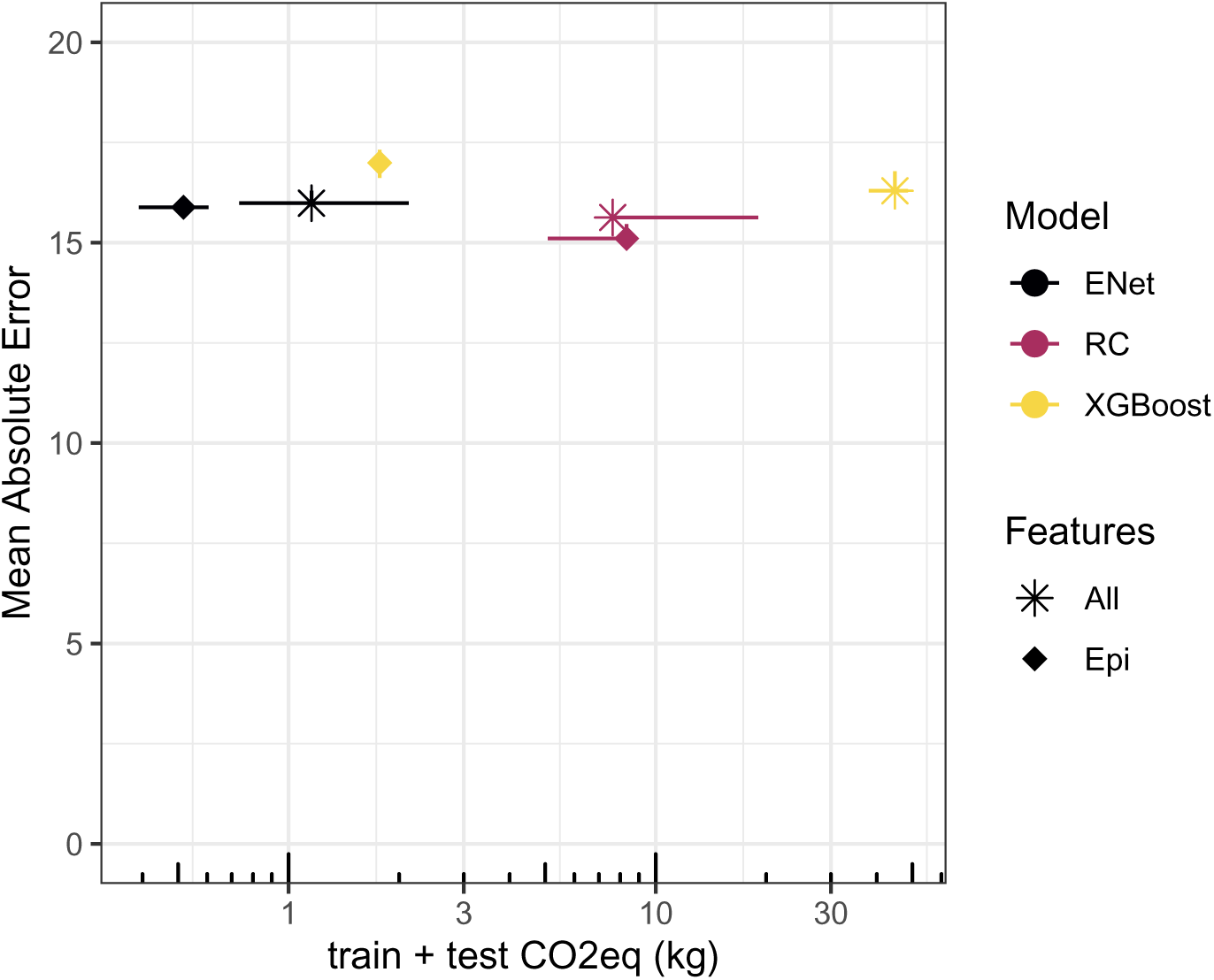
Carbon footprint and performance. Models were initiated on 202103-01 with monthly hyperparameter updates. Dots correspond to the median of three replications of the analysis; bars correspond to the minimum and maximum of the three replications.

### 6.4 Individuals

Implementation relied on the knowledge, attitudes, and involvement of key individuals. Interdisciplinary collaboration across informatics, epidemiology, biostatistics, and machine learning was essential (11–14) . Strong leadership and early engagement of the informatics team improved understanding of both data infrastructure and model assumptions.

Close collaboration between those developing, deploying, and interpreting the model helped ensure that forecasts were used appropriately. As shown in Figures 2 and 3, predictions are inherently uncertain and must be contextualized with other epidemiological indicators.

Finally, introducing more complex methods like RC highlights a potential barrier: these models are less familiar compared to linear regression. Addressing this gap is part of the motivation for this work. By documenting our experience, we aim to support knowledge sharing and reduce this barrier for future implementations.

### 6.5 Implementation process

As noted in Section 7.4, effective forecasting in routine practice requires close collaboration across multiple teams. This interdisciplinary work helps ensure that implementation is aligned with the realities of data acquisition, local constraints, and available infrastructure.

A staged approach is pragmatic: begin with simple models and selected features, then increase complexity as capacity grows. Enet with expert-selected variables provided a strong baseline (Figure 4), while RC offered modest improvements. This is further supported by Table 1, which shows that when limited information was available, none of the models substantially outperformed the baseline.

Finally, timely engagement of decision-makers through clear communication remains essential, as further discussed in Section 8.

## 7 Key Point 6: Communicate Effectively with Policymakers

Even the most accurate forecasting algorithm is ineffective if it does not translate into actionable decisions. Therefore, model predictions should be designed to support policymakers in making informed choices. However, given the inherent uncertainties surrounding SARS-CoV-2 dynamics, it remains unclear how best to communicate these predictions.

In our setting, access to model outputs was restricted to public health experts, who interpreted the predictions alongside other epidemiological indicators. These interpretations were then discussed at each hospital Covid-19 task force meeting, together with hospital indicators. This decision was primarily driven by two factors: (1) the moderate predictive performance of the models, which limited their standalone reliability, and (2) the rapid evolution of the epidemic, which was frequently influenced by external interventions such as lockdowns and curfews. Given these challenges, presenting raw model outputs to decision-makers without adequate context risked misinterpretation. Alternative communication strategies could be considered, but it is likely that model predictions should always be contextualized before being shared with policymakers to ensure proper understanding.

## 8 Discussion

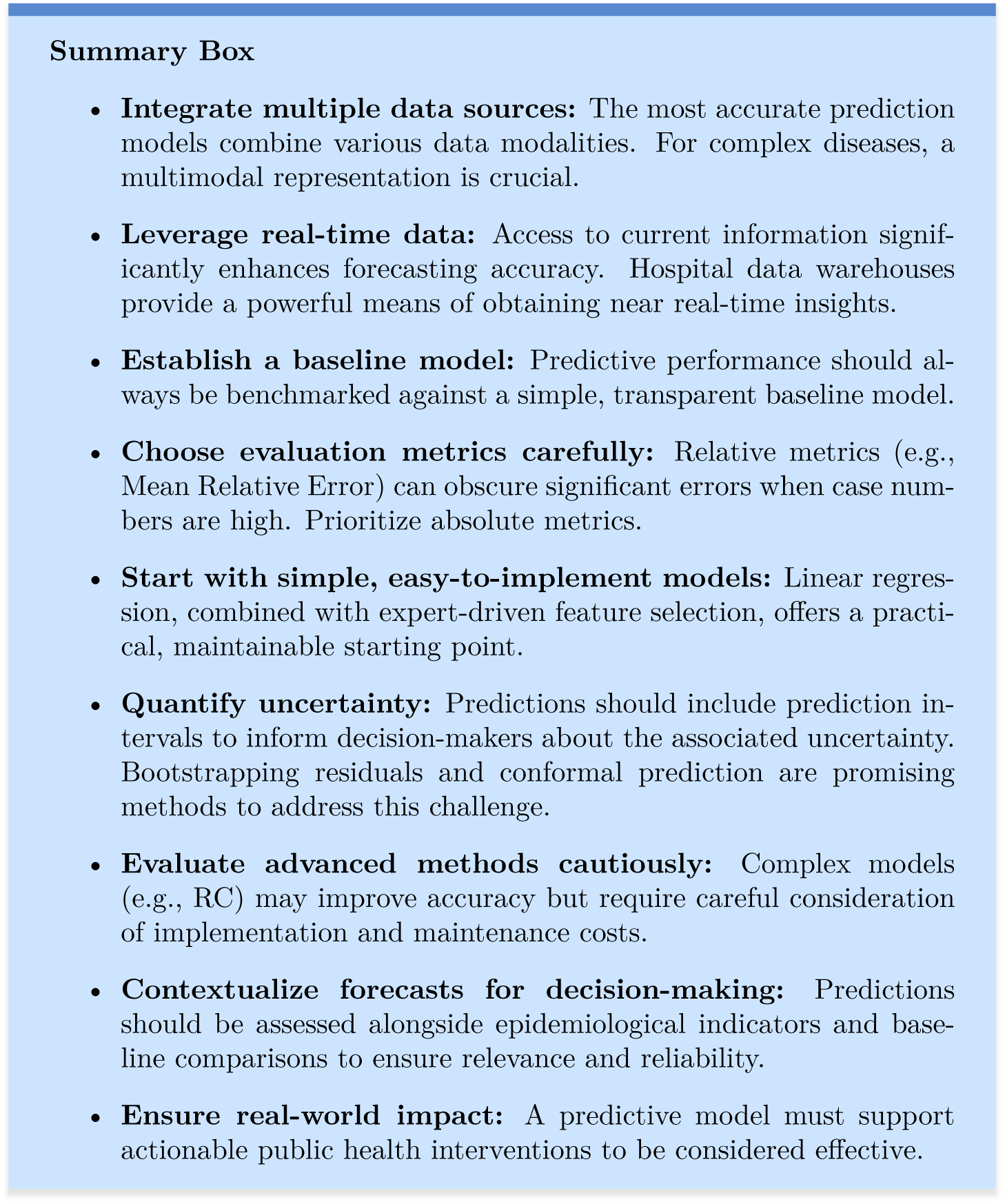

A summary of the main findings is provided in Box 9. This study of epidemic forecasting at BUH underscores the value of high-quality, real-time data from Hospital Data Warehouses and of a transparent baseline model to assess more complex approaches. Linear regression with expert-selected features offered a strong initial choice due to its simplicity, interpretability, and low maintenance, while models like RC with genetic algorithms improved accuracy but demanded greater computational resources, expertise, and upkeep. Forecasts should always be contextualized with epidemiological indicators and validated against baselines before informing policy, particularly in dynamic situations such as COVID-19.

Our findings align with recent work emphasizing diverse, real-time data integration and strong collaboration between modelers and decision-makers. Nunes et al. (10) advocate multimodal inputs such as wastewater, mobility, and social media data, highlighting challenges in trend shifts and the potential of ensemble methods. Endres-Dighe et al. (61) emphasize practical issues including computational demands, automation, and structured communication via dashboards and protocols. Across approaches, shared challenges were responsiveness to evolving data, automation, and stakeholder engagement.

Future work should improve adaptation to emerging epidemics. The cumulative validation framework used here weights all data points equally, which diminishes the influence of recent observations as the dataset grows. Approaches proposed by Montavon et al. (62) and Luksevcius et al. (58) suggest emphasizing recent or critical points, such as trend shifts.

Finally, black-box models like RC and XGBoost may reduce trust; explainable AI methods (e.g., Shapley values) can clarify which features drive predictions (63, 64).

Effective epidemic forecasting requires integrating diverse real-time data, grounding predictions in simple baselines, and choosing appropriate evaluation metrics. Starting with simple models enables rapid deployment while providing time to develop more complex approaches. Above all, forecasts must be contextualized and aligned with public health decision-making to achieve real-world impact.

## Acknowledgements

Experiments presented in this paper were conducted using the PlaFRIM experimental testbed, supported by Inria, CNRS (LABRI and IMB), Université de Bordeaux, Bordeaux INP and Conseil Ŕegional d’Aquitaine (see https://www.plafrim.fr), as well as by the MCIA (Mésocentre de Calcul Intensif Aquitain).

## Data availability

The data supporting this study is publicly available (65) . The code used in this study is also publicly available (66) .

## CRediT

RT, BH, and VJ contributed to conceptualization. VJ and RG were responsible for data curation. TF performed the formal analysis. Methodology was developed by TF, VT, XH, PL, DD, RG, VJ, BH, and RT. Project administration was carried out by RT and VJ. TF and DD developed the software, while RT, BH, VJ, and XH provided supervision. Visualization was conducted by TF. The original draft was written by TF, VT, VJ, BH, and RT, and XH, PL, DD, and RG contributed to writing—review and editing.

## 9 Supplementary

### 9.1 Supplementary methods

To illustrate various aspects of epidemic forecasting implementation, we use the example of SARS-CoV-2 forecasting at BUH, employing data and methods similar to those in Ferté et al. (12) . The following sections will detail the data, evaluation framework, modeling approaches, hyperparameter optimization and feature selection methods.

#### 9.1.1 Data

The objective of this study was to forecast the number of SARS-CoV-2 hospitalized patients at 14 days (ht+14) at BUH. To achieve this, we integrated data from multiple sources: department-level COVID-19 statistics from Santé Publique France (15), department-level weather data from the National Oceanic and Atmospheric Administration (NOAA) (16), and hospital-level COVID-19 data from BUH’s Electronic Health Records (EHR) (17) . The dataset covered the period from May 16, 2020, to January 17, 2022, resulting in a highdimensional problem with 409 predictors over 586 days, where each observation corresponds to a single day.

For preprocessing, we computed the first and second derivatives over the previous seven days to enrich the model with temporal dynamics. Additionally, we applied local polynomial regression smoothing with a 21-day span to reduce daily noise variations, following recommendations from Ferté et al. (11) . Finally, all features were scaled between -1 and 1 using maximum absolute scaling, where each value was divided by the maximum absolute value of the respective feature.

All datasets used in this study are publicly available. Weather data can be accessed via Smith et al. (16) through the R package worldmet (67), and vaccine data can be downloaded from Etalab (15) here. EHR data are available on Dryad (17), with privacy protections in place. For that purpose, EHR values below 10 patients were obfuscated to 0. To account for this, when evaluating model performance, outcomes, forecasts, and hospitalization values of 0 were replaced with 10. The full dataset and code are accessible via a GitHub repository https://github.com/thomasferte/Ferte2024ICMLHighDimensionReservoir.

#### 9.1.2 Evaluation framework

Model performance was evaluated using two different implementation periods: an ”early” model starting on September 2, 2021, and a ”late” model starting on March 1, 2021. Data prior to these start dates (i.e., before September 2, 2021, for the early model and before March 1, 2021, for the late model) were used solely for identifying relevant hyperparameters. The remaining data were used to assess model performance, with hyperparameters being updated (or left unchanged) on a monthly basis.

These two periods were chosen to examine how model performance differs when trained with limited historical data versus when more information is available. The early period represents a scenario where forecasting is more challenging due to limited prior observations, while the late period illustrates what can be achieved with a longer historical context. Our hypothesis was that, with limited data, simpler models incorporating expert-driven feature selection would outperform more complex approaches. However, as more information becomes available, more sophisticated models might demonstrate superior performance. The primary evaluation metric was MAE, while secondary metrics included Median Relative Error (MRE), Mean Absolute Error to Baseline (MAEB), and Median Relative Error to Baseline (MREB). These metrics were defined as fol-lows:

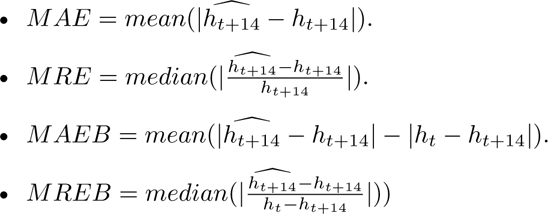

The relevance of these different metrics will be discussed in the following sections, particularly by contrasting absolute and relative errors and emphasizing the importance of baseline-derived metrics.

To limit computational costs, models were trained and evaluated every other day rather than on a daily basis using an accumulative validation scheme (58). This scheme, illustrated in Figure S1, updates hyperparameters monthly based on all data available up to the beginning of each month, as described in Section 10.1.4. The optimal set of hyperparameters is selected and used to forecast the following month. At the start of the next month, new candidate hyperparameters are evaluated on all available data up to that point, and the best set is again chosen for forecasting. This cycle continues until the end of the dataset.

#### 9.1.3 Models and Implementation

We compared three different models: ENet, XGBoost, and RC.

- ENet was selected as a representative of statistical models. It is a simple, interpretable model that is easy to use and implement.
- XGBoost was chosen as a machine learning model. As a tree-based algorithm, it has been widely applied in time series forecasting, including SARS-CoV-2 forecasting.
- RC was selected as a neural network approach. Originally introduced by Jaeger et al. (19) and Maass et al. (20), RC is a distinct machine learning paradigm tailored for processing dynamic time series data (21). Unlike conventional recurrent neural networks (RNNs), RC consists of a randomly initialized recurrent layer (the reservoir), where only the output layer (read-out) is trained. This structure enables non-linear transformation of inputs into a high-dimensional space, capturing complex temporal dependencies while reducing computational cost compared to traditional RNNs (22). RC has been applied in various fields, including bird song analysis, language processing, epidemic modeling, power plant monitoring, internet traffic forecasting, and financial markets (24–28). However, selecting appropriate RC hyperparameters remains a challenge due to their impact on reservoir dynamics. To address this, we adopted the Genetic Algorithm (GA) optimization approach proposed by Ferté et al. (12) .

Given the poor performance of heavier models such as Transformers in this context (12), they were not included in this study. The RC architecture used in this work featured 500 recurrent units, with both the input layer and reservoir connected to the output layer. Hyperparameter optimization was performed by evaluating the MAE of the median forecast across three RC instances with the same hyperparameter set. To assess the impact of hyperparameter optimization, we conducted an additional analysis using 20 RC instances instead of three.

For forecast evaluation, we computed the median forecast from 40 RC instances, each corresponding to one of the top 40 hyperparameter sets. Given the stochastic nature of RC due to randomly initialized reservoir connections, aggregating multiple forecasts helps improve stability and robustness.

#### 9.1.4 Hyperparameter Optimization and Model Training

Hyperparameter selection was performed using random search for XGBoost and ENet and Genetic Algorithm (GA) for RC, as proposed by Ferté et al. (12) . At model initialization, approximately 3,500 hyperparameter sets were evaluated, with 1,500 new evaluations added each month. Details on hyperparameter ranges are provided in Supplementary Table 4. The optimization process targeted:

- All hyperparameters of ENet
- Key hyperparameters of XGBoost, as identified by Yang et al. (68)
- Main hyperparameters of RC, based on the ReservoirPy documentation (69)

Following Ferté et al. (12), models were trained to forecast hospitalization variation rather than absolute hospitalization counts. However, all performance metrics and visual representations were reported on the raw hospitalization scale for interpretability.

To account for the non-stationary nature of COVID-19 time series data, we used a sliding window of one year for training. This approach was found to improve forecasting performance in previous research (12) .

#### 9.1.5 Feature selection

To explore the potential improvement in forecasting by incorporating expert knowledge, we developed a feature selection approach. Initially, all 409 predictors were included as input features. Feature selection was then performed using different methods depending on the model:

- For RC, feature selection was carried out via Genetic Algorithm (GA).
- For ENet, feature selection was embedded directly through L1 penalization.
- For XGBoost, feature selection occurs inherently as the algorithm selects relevant features during the tree-building process.
- Additionally, we proposed selecting a subset of features a priori, based on expert knowledge. The chosen features were:
- Hospitalization data
- Number of positive RT-PCR tests
- Proportion of positive RT-PCR tests among individuals aged 60 and older
- IPTCC, a weather index specifically developed for SARS-CoV-2 forecasting
- Cumulative number of first-dose vaccinated individuals
- Number of people presenting to the emergency department with ”COVID19” mentioned in their Electronic Health Records (EHR)

Our hypothesis was that incorporating expert-driven feature selection would improve forecast accuracy by leveraging external information to better distinguish signal from noise. We expected this effect to be particularly pronounced in the early phase of the epidemic when limited data was available, and the signal-noise ratio is low.

### 9.2 Supplementary uncertainty quantification

**Fig. S1:**
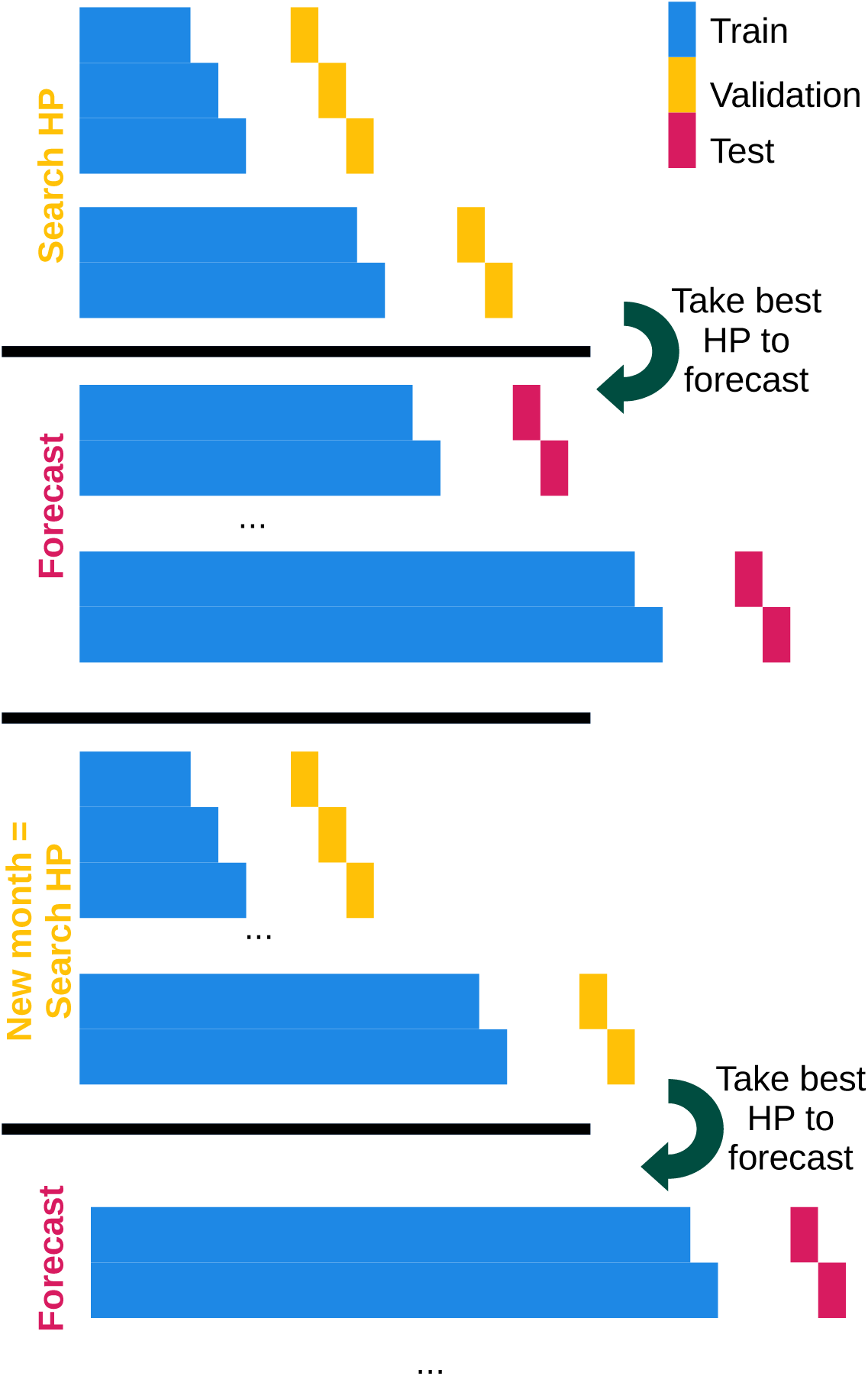
Cross-validation scheme.

**Fig. S2:**
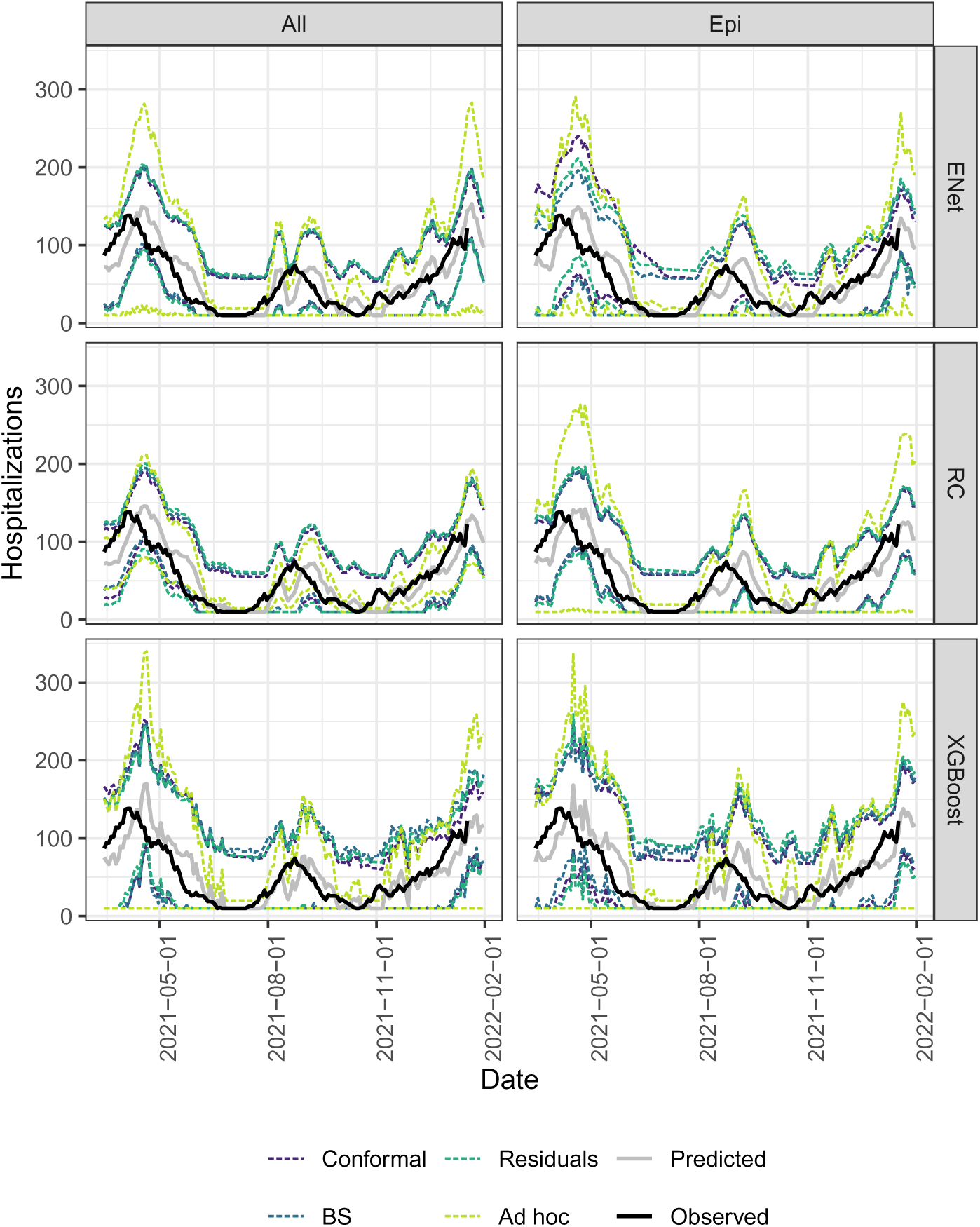
Prediction intervals of Residuals, Bootstrap (BS), AgACI conformal inference (Conformal) and Ad hoc methods.

